# Does one session of dry needling effect vital capacity in people with Parkinsons Disease?

**DOI:** 10.1101/2023.03.14.23287205

**Authors:** Ariany Klein Tahara, Ada Clarice Gastaldi, Abel Gonçalves Chinaglia, Rafael Luiz Martins Monteiro, Vitor Tumas, Paulo Roberto Pereira Santiago

## Abstract

Respiratory function problems are caused by both motor and non-motor symptoms of Parkinson’s disease (PD). One major problem involving the changes in respiratory function in people with PD is a decrease in chest movement caused by musculoskeletal problems around the neck and upper trunk. The relationship between these respiratory changes and pulmonary volume in people with PD can lead to the main cause of mortality as the disease progresses. Dry needling technique (DNT) is a myofascial release technique that has been reported to provide an immediate effect on pain, decreased muscle spasm or tightness and lead to restoration of range of motion of upper trunk by using needles to stimulate hypersensitive and painful nodules in the musculature called trigger points (TP). However, to date, the use of this technique has not been reported to release muscle tightness or pain in people with PD. The present study aimed to explore the effects of a single session of bilateral DNT to the trapezius muscles trigger point on chest expansion and vital capacity which could lead to improvements in respiratory function in people with PD. Thirty-eight people with PD were randomly allocated to DN and Sham needling treatment groups. The maximum and mean volumes of vital capacity were assessed by using a ventilometer at pre-intervention, immediately after intervention, and one-week follow-up. Mixed Model Analysis of Variance (MM ANOVA) tests with post hoc pairwise comparisons were used to test the differences between groups and assessment time points. No interaction effects were found between groups and assessment time points for maximum and mean vital capacity volume. In addition, no statistically significant main effects of DNT were found for both groups and assessment time points for either maximum or mean vital capacity volume. These findings can provide evidence that a single session of dry needling does not help to improve respiratory function in people with PD. However, slight improvements in vital capacity were observed in the DN group, which may be clinically relevant when considering progressive neurodegenerative disease. More sessions of dry needling need to be explored over a greater time period to determine if longer term effect are possible with this treatment.

## INTRODUCTION

Parkinsons disease (PD) is the second most prevalent neurodegenerative disease in the elderly over the age of 60, second only to Alzheimers disease. Its pathophysiology involves the degeneration of dopaminergic neurons in the substantia nigra with consequent decrease in dopamine due to alterations in the nigrostriatal pathways or genetic mutations^1^.

PD mostly affects male adults over 60 years of age, with changes in motor skills in different degrees, as well as their progression, cognitive impairment and response to treatment being different in each individual^2,3^. It is a disease primarily known and stigmatized by the motor repercussions, with the main signs and symptoms including; resting tremor, bradykinesia with plastic appendicular stiffness and postural changes, and balance and gait changes including freezing or scrambled gait^4–6^. In addition to motor repercussions caused by PD, there are also changes in lung function, such as decreases in the amplitude of the chest and lung volumes that are associated with increased spasm and stiffness of respiratory muscles which can be observed even in the initial stages of the disease^7,8^. Pulmonary compliance decreases due to a number of factors including limitations in trunk extension and a reduction of the articular amplitude of the thorax and spine, secondary to arthrosis and other thoracic structural changes such as increased kyphosis^9^. The physiological ageing of the respiratory system includes loss of elasticity, reduction of neural stimulation of the respiratory musculature, dilation of the alveoli and alterations in the flow, volumes and capacities, all of which have repercussions on the respiratory mechanics of inspiration and exhalation. These ventilation-related changes in individuals with PD in advanced stages of the disease can generate accumulation of purulent which can lead to the onset of infectious conditions, pneumonia aspiration or pulmonary embolism, which are the main causes of mortality of these neurodegenerative diseases^10,11^.

Trigger points (TP) are hypersensitive and palpable nodules which are easily identified and are formed through histological changes such as decreases in the size of the sarcomeres or tissue hypoxia, and biochemical changes such as excess of acetylcholine and substance P that influence the sensitisation of the central and peripheral nervous system. The presence of TP in muscles or connective tissues is called Myofascial Pain Syndrome (MPS) and is commonly found in patients with muscle aches^12–14^, and is associated with pain, spasms, stiffness, muscle weakness, decreased range of motion (ROM) and autonomic dysfunction^13^.Dry needling technique (DNT) is an intervention that uses filiform needles as physical agents to penetrate the skin, reaching and stimulating trigger points within muscles and connective tissues that are usually associated with musculoskeletal pain and movement disorders^12,15^.The application of dry needling has shown an immediate effect on increasing the pressure pain threshold and ROM, decrease muscle tone and pain, and has been widely used in the clinical environment to address a variety of musculoskeletal conditions as it is simple to perform, minimally invasive, low cost and has been reported to have good effectiveness^12,16–18^.

It is known that pulmonary impairment may be present already in the early stages of PD, with abnormalities in lung volume levels^19^. In addition, muscle spasms and stiffness around the head and neck area in people with PD can affect the whole body, which can effect respiratory mechanics by reducing the pulmonary volumes due to lower chest expansion capacity^7,9^. This combination between muscle spasm and lower chest expansion may alter the posture in this population, with an adoption of a stooped posture typical of people with PD. These factors might be as a result of the presence of TP formed from structural and biochemical changes in muscle tissue, specifically on accessory breathing muscles which tend to restrict the movement of the extension and expansion of the chest^12,20^.

The aim of this study was to release the trigger point of trapezius muscles in people with PD using the dry needling intervention. We hypothesized that releasing spasms and pain within the trapezius muscle may help to improve respiratory function in people with PD through increases in vital capacity measured using a ventilometer.

## MATERIALS AND METHODS

### Patients

This longitudinal, prospective, concurrent and uncontrolled study design was investigated in people with PD which were being diagnosed and monitored by a neurologist at the Neurology Service of Clinics Hospital of Medical School of Ribeirão PretoUniversity of São Paulo (FMRP-USP). For the sample calculation we used the statistical software G*Power (ver. 3.1.9.7; Heinrich-Heine-Universität Düsseldorf, Düsseldorf, Germany) and calculated from a previous study of DNT on PD population^21^. The sample size calculation was reported that 14 participants per group were required for each group, 28 participants in total. Based on a statistical power of 95% at a 5% significance level. Taking into consideration at least 20% drop-out, therefore, 38 participants were recruited to this study. Participants were randomized into two groups, a Dry Needling group (DN) and the Sham Needling by a random allocation sequence generated using the Research Randomizer website (www.randomizer.org). This study was approved by the Ethical Committee of School do Physical Education and Sport of Ribeirão Preto/University of São Paulo. All participants read and signed the consent form in agreement with the National Ethics Committee in line with the Declaration of Helsinki before data collection started.

### Inclusion criteria

Patients were recruited if they were over 50 years old, had a clinical diagnosis of idiopathic PD according to the Hughes criteria (London Brain Bank)^2,22^ had mild to moderate PD using a modified Hoehn and Yahr scale ranging the score of 1.5 to 3.0, using levodopa as PD medication, were able to walk and remain in the orthostatic position, were able to respond to commands, and had a TP on the trapezius muscles. All assessments and treatments were carried out in the on treatment state at least 60 minutes after taking medication.

### Exclusion criteria

Individuals using auxiliary devices for walking who had pulmonary involvement (i.e., chronic obstructive pulmonary disease (COPD) and asthma), smokers, a history of thoracic surgery, dyskinesias induced by levodopa in more than 50% of the wake period, a diagnosis of associated dementia syndrome, and any neurological deficit that prevented them from understanding or attending to commands were excluded. Furthermore, individuals were excluded if they reported a contraindication to dry needling, including; a patient with needle fobia, patient unwillingness (fear, patient belief) to undergo dry needling, age-related factors, or medical emergency or acute medical condition, e.g., occlusion and coagulopathy, and patient with lymphedema.

### Evaluation

#### Unified Parkisons Disease Rating Scale (UPDRS)

People with PD in both groups were assessed the demographic data and PD severity using Unified Parkisons Disease Rating Scale (UPDRS) - part III motor assessment. In addition, the evaluations of vital capacity and lung volumes were assessed by using the ventilometer^23^ at three time points: 1) pre-intervention assessment as baseline (BL); 2) post-intervention assessment in both DN or Sham groups as immediately after the intervention (IA) and 3) follow-up after 1-week of intervention (FU).

The UPDRS considers four main items: mental activity, behavior, and humor; activities of daily living (at different stages: ON and OFF); motor examination; and complications of drug therapy. The total score is 199, and each question has a score ranging from 0 (zero) to 4 (four), where 0 indicates no involvement, 1 indicates a detectable disorder, 2 indicates a moderate disorder, 3 indicates a significant disorder, and 4 indicates no function or a severe disorder. The lower the score, the lower the PD impairment^3,24^. This study reported only the results of the motor examination at the initial evaluation to demonstrate the severity of participants in each group.

#### Ventilometer

Lung volume and capacity were measured using a ventilometer, as the spirometry, it is the most useful technique to diagnose pulmonarys mechanical limitations and respiratory functions. It consisted of measures of the vital capacity, which comprises the maximum amount of air exhaled from the lungs after a maximum inhalation. An analogic ventilometer Wright Mark 8 (nSpire Health Inc., Longmont, CO, US) and a disposable plastic mouthpiece was used for these measures. (Figure 1) The participant was seated, with a chair supporting the back. Then, the participant performed deep inhalation until total pulmonary capacity was reached. At this point, a nasal clip was placed on the participants nose to prevent air escape through the nose, and the participant was instructed to exhale continuously and slowly through the mouthpiece until feel the sensation of emptying of air from lungs, which meant reached the residual volume. The assessment was applied three times to measure the maximum and the mean volumes of these three times, with intervals of 60 seconds between them. The highest volume was considered^23,25,26^. However, concerning of demographic data, normalization of the maximum and mean of vital capacity were performed as percentage of values expected for controls of similar age, sex and height. In addition, there is a way to easily and continuously monitor the heart rate (HR) and the percentage of partial oxygen saturation (SpO2) that is present in the blood during the assess of the vital capacity with the ventilometer^27^.A pulse oxymeter G-Tech Oled Graph (Beijing Choice Electronic Technology Co., Ltd., Beijing, China) was placed on the distal phalanx of the participants index finger with the function of monitoring data while the participant performed deep inhalations and slow exhalations up to residual volume.

**Figure 1:**
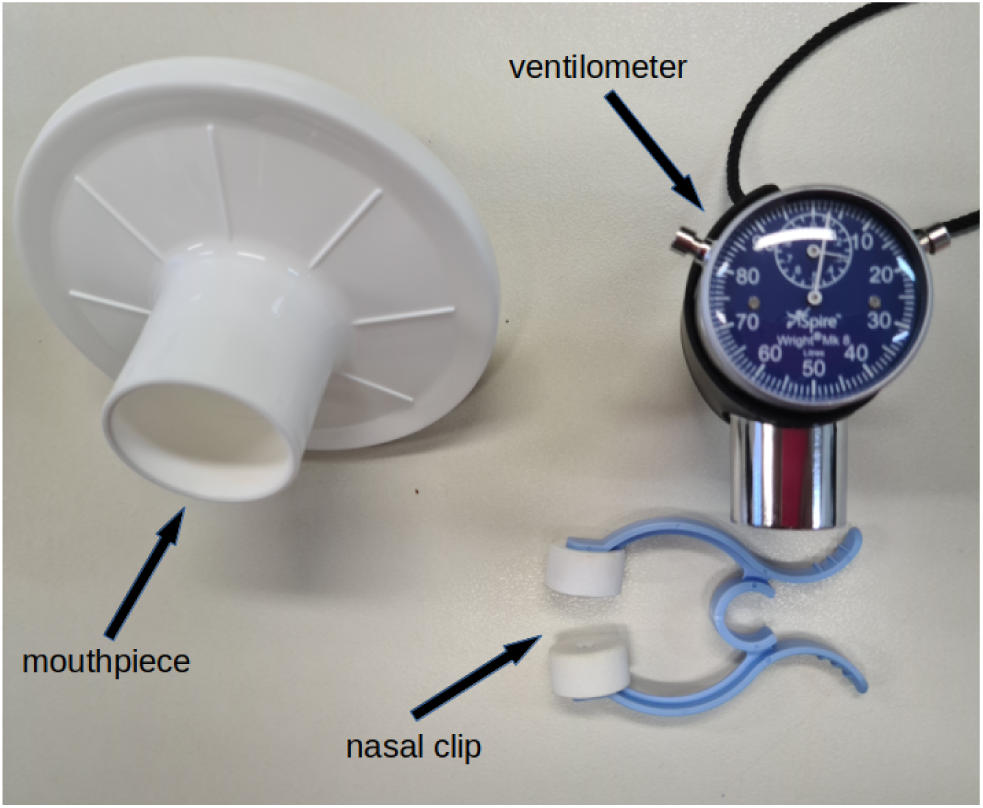
Equipment used to assess lung volumes and capacities in the study.

#### Dry Needling (DN) intervention and Sham Dry Needling (Sham) with esthesiometer

As mentioned above, the participants were randomized into two groups (DN or Sham groups). Prior to the intervention, the participant was asked to sit comfortably with their back support on the chair, hands on thighs, and feet on the floor. Alcohol (70%) was used in the trapezius muscles, the most affected by TP on neck-shoulder muscles in people with mechanical neck pain^28^, bilaterally to remove oil and clear the skin before the intervention. For the DN, we inserted filiform needles (0.25×40mm) to penetrate the skin, subcutaneous tissues, and muscle with the intent to mechanically disrupt tissue in a fast-in and fast-out technique (Figure 2A)^29^. There was no time limit for fast-in and fast-out movements; it was completed when the physical therapist noticed a reduction in the size of the TP or its complete disappearance. In addition, for the Sham group, we include a phase in the intervention, where we replace the needle with the thickest filament of the esthesiometer to have a consistent resistance to simulate movement of the needle and without deformation of the filament (magenta filament 1 × 15 mm) to simulate the DN (Figure 2B)^30^. The intervention was performed without the participant being able to see the needle or the esthesiometer. All throughout the entire intervention, which lasted 30 minutes for each patient individually in each group, no incidents or complications were reported in either group.

**Figure 2:**
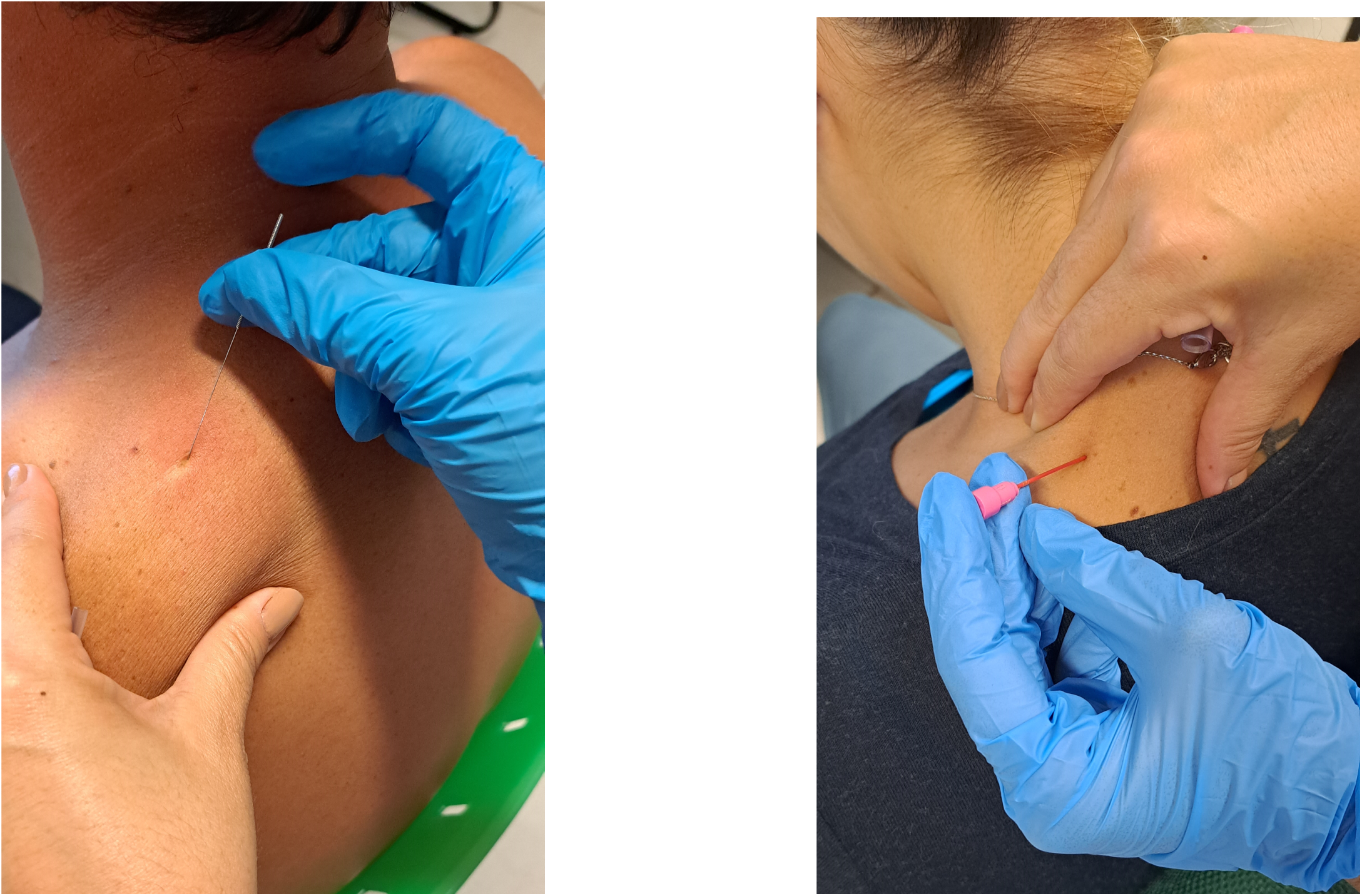
Two forms of intervention in the Upper Trapezius: Dry Needling and sham with the esthesiometer.

### Statistical analysis

The distribution of the data was tested using Shapiro-Wilk tests and all variables were found to be suitable for parametric testing. To compare participant characteristics between the groups independent t-tests were performed. The factors included the two groups (DN and Sham) and three time points (baseline, immediately after and 1-week follow-up) which were explored using a Mixed Model Analysis of Variance (MM ANOVA) to determine any significant interactions between the factors or main effects. The statistical significance level was set at *p* <0.05, and SPSS for Windows, Version 18.0 (Chicago SPSS Inc.) was used for all analysis.

## RESULTS

Thirty-eight patients were invited to participate in this study. However, one individual from the DN group did not showed up on follow-up assessment. Therefore, thirty-seven patients in total (DN = 18 and Sham = 19) were included in the analysis. There were no significant differences between groups with regard to age, height, weight and body mass index underlying disease which was tested using independent t-tests (*p* > 0.05). Only the stage of PD shows significant differences (*p* = 0.008) between groups (Table 1). There were no interactions between groups and assessment time points for the maximum volume (*p* = 0.852) and the mean volume (*p* = 0.867) (Table 2). In addition, no main effects of DNT for both group (*p* = 0.301 and *p* = 0.572 for the maximum and mean volumes, respectively), and between BL and IA assessments (*p* = 0.776 and *p* = 0.698 for the maximum and mean volumes, respectively), BL assessment and FU (*p* = 0.573 and *p* = 0.607 for the maximum and mean volumes, respectively) and IA assessment and FU (*p* = 0.796 and *p* = 0.918 for the maximum and mean volumes, respectively) were found for both maximum and mean expiratory volumes (Table 3).

**Table 1:**
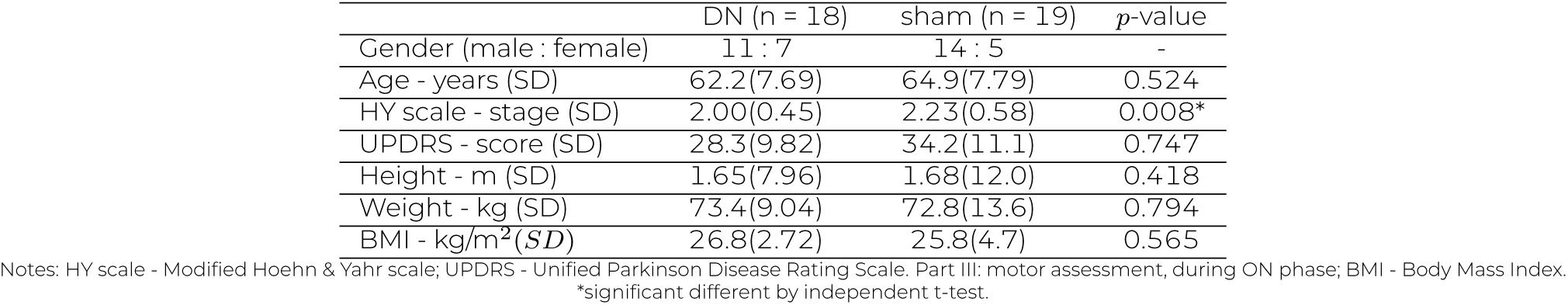
Summary of patients’ demographics, reporting mean (standard deviations) for the two groups dry needling (DN), sham and their corresponding *p*-values.

|  | DN (n = 18) | sham (n = 19) | *p*-value |
| --- | --- | --- | --- |
| Gender (male : female) | 11 : 7 | 14 : 5 | - |
| Age - years (SD) | 62.2(7.69) | 64.9(7.79) | 0.524 |
| HY scale - stage (SD) | 2.00(0.45) | 2.23(0.58) | 0.008* |
| UPDRS - score (SD) | 28.3(9.82) | 34.2(11.1) | 0.747 |
| Height - m (SD) | 1.65(7.96) | 1.68(12.0) | 0.418 |
| Weight - kg (SD) | 73.4(9.04) | 72.8(13.6) | 0.794 |
| BMI - kg/m<sup>2</sup> (SD) | 26.8(2.72) | 25.8(4.7) | 0.565 |

Notes: HY scale - Modified Hoehn & Yahr scale; UPDRS - Unified Parkinson Disease Rating Scale. Part III: motor assessment, during ON phase; BMI - Body Mass Index. \*significant different by independent t-test.

**Table 2:**
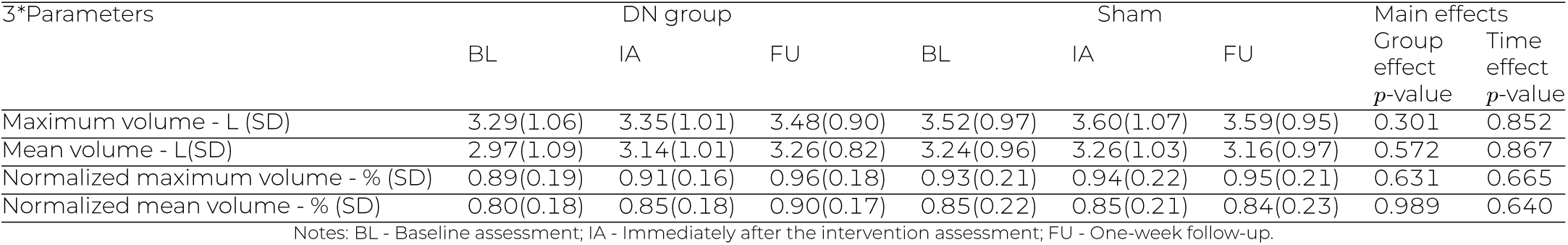
Results of Mix Model ANOVA.

| Parameters | DN group |  |  | Sham |  |  | Main effects |  |
| --- | --- | --- | --- | --- | --- | --- | --- | --- |
|  | BL | IA | FU | BL | IA | FU | Group effect<br>*p*-value | Time effect<br>*p*-value |
| Maximum volume - L (SD) | 3.29(1.06) | 3.35(1.01) | 3.48(0.90) | 3.52(0.97) | 3.60(1.07) | 3.59(0.95) | 0.301 | 0.852 |
| Mean volume - L(SD) | 2.97(1.09) | 3.14(1.01) | 3.26(0.82) | 3.24(0.96) | 3.26(1.03) | 3.16(0.97) | 0.572 | 0.867 |
| Normalized maximum volume - % (SD) | 0.89(0.19) | 0.91(0.16) | 0.96(0.18) | 0.93(0.21) | 0.94(0.22) | 0.95(0.21) | 0.631 | 0.665 |
| Normalized mean volume - % (SD) | 0.80(0.18) | 0.85(0.18) | 0.90(0.17) | 0.85(0.22) | 0.85(0.21) | 0.84(0.23) | 0.989 | 0.640 |

Notes: BL - Baseline assessment; IA - Immediately after the intervention assessment; FU - One-week follow-up.

**Table 3:** Post hoc comparisons for the main effects of groups and assessment time points as shown by the mixed model analysis of variance, where no interactions were indicated.

| Variables | Comparison | Mean Diff<br>(SE) | *p*-Value | CI of Diffs |  |
| --- | --- | --- | --- | --- | --- |
|  |  |  |  | Lower<br>Bound | Upper<br>Bound |
| Maximum volume (L) | BL vs IA | -0.07 (0.24) | 0.776 | -0.549 | 0.411 |
|  | BL vs FU | -0.13 (0.22) | 0.573 | -0.582 | 0.325 |
|  | IA vs FU | -0.06 (0.23) | 0.796 | -0.520 | 0.400 |
|  | DN to Sham | 0.2 (4.17) | 0.301 | -0.179 | 0.575 |
| Mean volume (L) | BL vs IA | -0.9 (0.24) | 0.698 | -0.571 | 0.384 |
|  | BL vs FU | -0.12 (0.23) | 0.607 | -0.567 | 0.334 |
|  | IA vs FU | -0.02 (0.01) | 0.918 | -0.473 | 0.426 |
|  | DN to Sham | 0.11 (0.19) | 0.572 | -0.266 | 0.480 |
| Normalized maximum<br>volume (%) | BL vs IA | -0.01 (0.05) | 0.770 | -0.108 | 0.080 |
|  | BL vs FU | -0.04 (0.05) | 0.378 | -0.136 | 0.052 |
|  | IA vs FU | -0.03 (0.05) | 0.549 | -0.121 | 0.065 |
|  | DN to Sham | 0.02 (0.04) | 0.632 | -0.058 | 0.095 |
| Normalized mean<br>volume (%) | BL vs IA | -0.21 (0.05) | 0.651 | -0.114 | 0.072 |
|  | BL vs FU | -0.05 (0.05) | 0.346 | -0.142 | 0.05 |
|  | IA vs FU | -0.03 (0.05) | 0.606 | -0.119 | 0.070 |
|  | DN to Sham | -0.001 (0.04) | 0.989 | -0.077 | 0.076 |

Notes: L - Liters; Diff: different; CI: Confident Interval; % - percent; DN: Dry needling group; Sham: Sham needling group; BL: Baseline assessment; IA: Immediately after the technique assessment; FU: one-week follow-up; DN: Dry needling group; Sham: Sham needling group

No interaction between group and assessment time points for the normalized maximum volume (p = 0.665) and normalized mean expiratory volume (*p* = 0.640) (Table 2). Furthermore, there was no significant main effect of DNT for both groups (*p* = 0.631 and *p* = 0.989 for the normalized maximum and normalized mean volumes, respectively), and between BL and IA assessments *p* = 0.770 and *p* = 0.651 for the normalized maximum and normalized mean volumes, respectively), BL assessment and FU *p* = 0.378 and *p* = 0.346 for the maximum and mean volumes, respectively) and IA assessment and FU *p* = 0.549 and *p* = 0.606 for the normalized maximum and normalized mean volumes normalized maximum and normalized mean volumes, respectively) were found for both the maximum and mean volumes (Table 3).

When we considered the raw data in Tables 2 and 3, this showed the maximum and mean volumes and the normalized maximum and normalized mean volumes were increased in the DN compared to the Sham and immediate effects of dry needling as in the immediately after intervention assessment were increase when compared to baseline assessment. The boxplot of lungs volumes (maximum and mean) and normalized maximum and mean (Figure 3) and graphics for Maximum and Mean volumes (Figures 4A,4B) and for Normalized Maximum and Mean volumes (5C,5D) below show this data.

**Figure 3:**
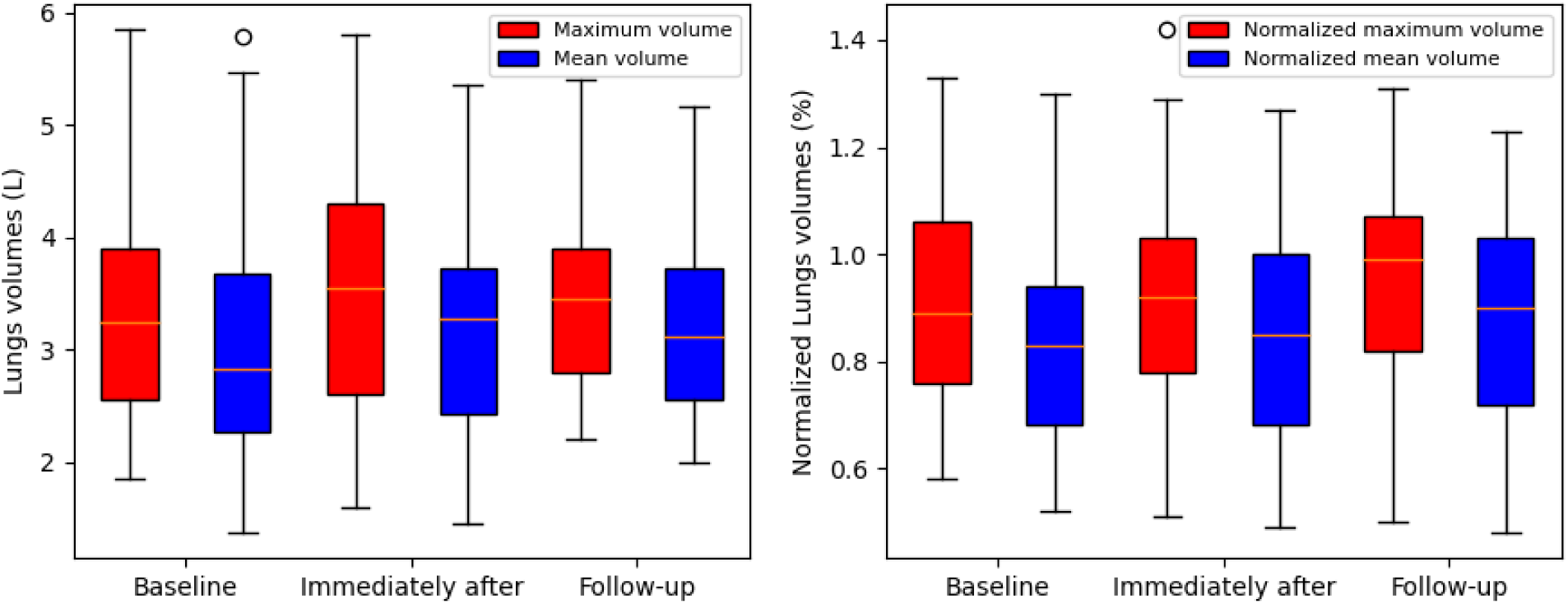
Boxplots of the variables: Lungs volume (maximum and mean volumes) in liters and Normalized maximum and mean values as a percentage. Both at baseline, immediately after intervention and follow-up.

**Figure 4:**
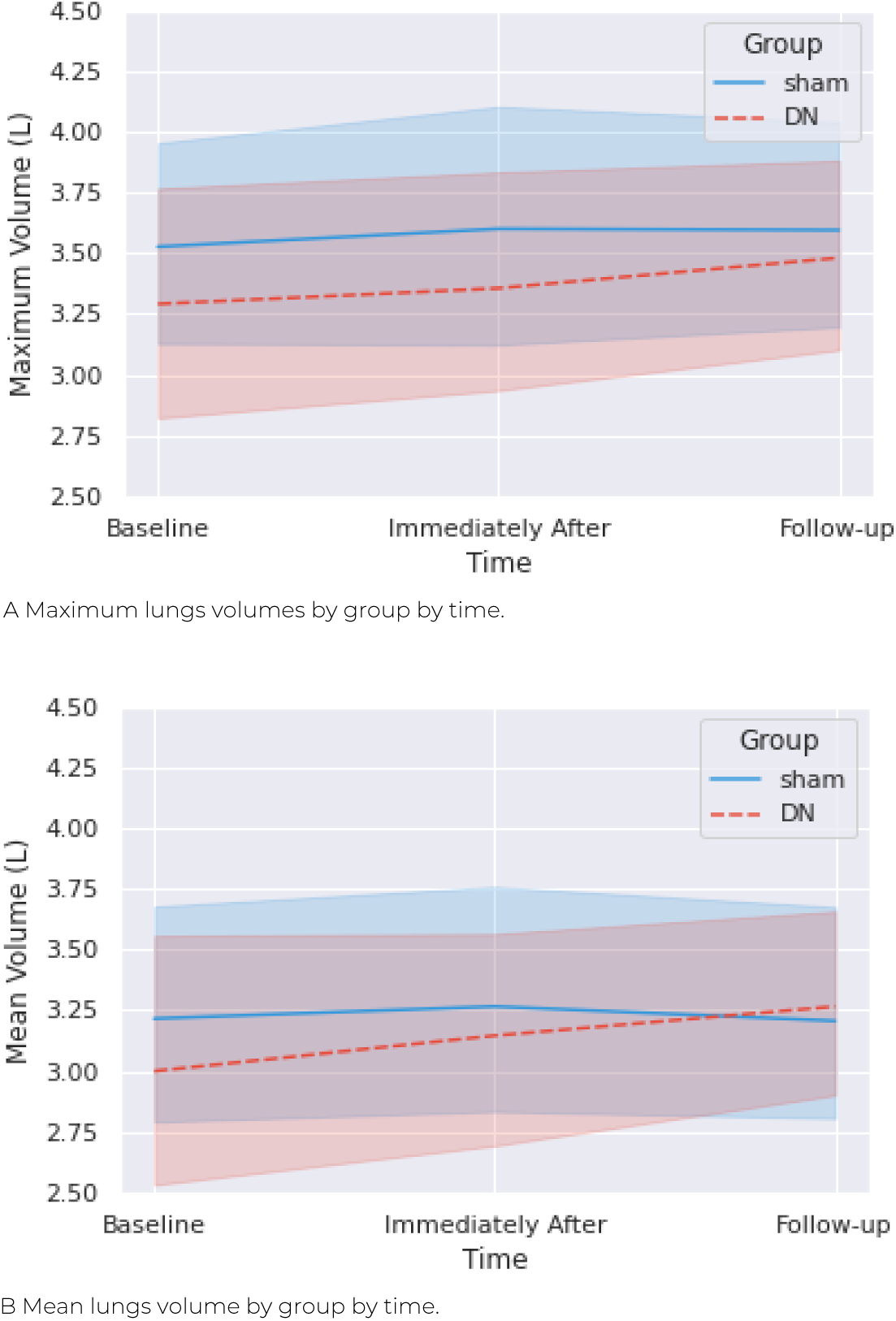
A) Maximum volumes of lungs by time of assessment and by group; B) Mean volumes of lungs by time of assessment and by group;

**Figure 5:**
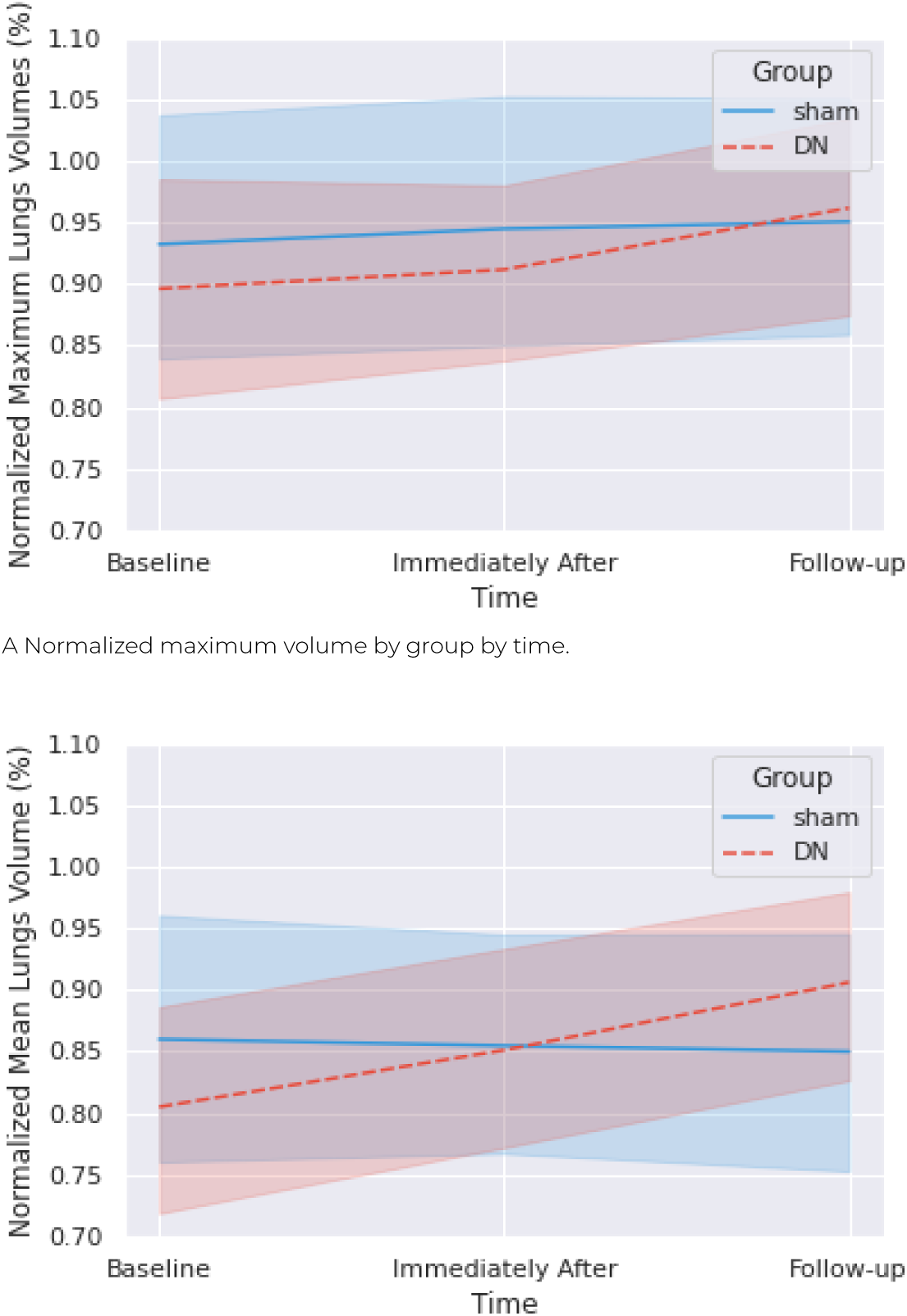
A) Normalized values of maximum volumes of lungs by time of assessment and by group; B) Normalized values of mean volumes of lungs by time of assessment and by group.

## DISCUSSION

This study is the first study that aimed to investigate the effects of one session of the dry needling technique to release TP on the muscles affected by PD symptoms and improve the vital capacity by lungs volume. The main finding was that the PD in the dry needling intervention was not statistically significant for all variables at three different time points. However, an increase in the mean raw data of maximum and mean volumes, as well as the normalized maximum and mean volumes, is seen in PD participants of the DN when compared between baseline and immediately after intervention assessment.

The role of DNT in the treatment of pain and stiffness for patients with TP in the trapezius has been suggested^16,31–33^. The release of myofascial TP by DNT have demonstrated good results if compared to sham needling immediately after the technique in short-term and midterm follow-ups concerning of pain and range of motion^34–36^. We found no evidence that DNT was being used in the PD population, despite the fact that they displayed symptoms such as rigidity, pain, and decreased range of motion. We are aware that such affections in PD occur due to the progressive nature of the disease, but considering how this can affect their routine, their daily living activities, and their tendency to increase their flexor posture and consequently tend to restrict their thoracic expansibility, and also taking into consideration the ease of use of the technique, its effectiveness in the general population, and the low cost, we chose to test it for a possible new non-pharmacological approach for PD.

Our sample consisted of twenty-five men and twelve women. These findings may corroborate previous research that found a higher prevalence of PD in men over the age of 60^2,22^. Our protocol of assessments and intervention was executed after one hour levodopa intake to ensure the period ON of all patients. Despite being the gold standard for treating motor disorders, there is debate about its role in treating respiratory dysfunction in PD, as Oliveira et al. (2022) stated that the effect of levodopa on respiratory function is not significant in PD patients at the mild stage^37,38^.

The review of Pokusa et al. (2020)^39^ is in line with authors who cited pulmonary complications of PD as the most frequent cause of death in this population^10,11,19,38,40^. This study also points out the impairment of the respiratory system even in the early stages of the illness, when the diagnosis tends to be complicated and masked by other symptoms. Our sample had patients with different times of diagnosis of PD, varying from two months to twenty-five years, and we could observe that the average normalized volume was around 8090% of lung capacity according to age, sex, and height, which do not indicate a ventilatory abnormality in PD. Regarding the maximum normalized volume, both groups had a slight increase in values over time of assessments, more noticeable in DN, which was not statistically significant, but based on the assumption of the progressive nature of PD and the worsening of lung involvement with the evolution of the disease, this modest increase may be useful for clinicians in an attempt to improve the limited lung volumes cited in some studies^39–41^.

The PD population usually does not have respiratory symptoms, although the pulmonary alterations are already installed, and this is mainly due to the motor impairments that make them prone to sedentary life^41,42^. Most of the people with PD in our sample reported that, while the interventions were being performed, they were active and independent in their daily lives, engaging in activities such as Pilates, swimming, hiking, and biking; this could explain the study’s findings, which showed no significant increase in pulmonary volumes, because a large portion of the sample was not sedentary. PD has obstructive and restrictive patterns. PD’s obstructive pattern affects the upper airways and may compromise health even in the asymptomatic phase of the disease, becoming more noticeable when there is a high degree of motor impairment caused by rigidity and fatigue in the thyroarytenoid muscles and causing speech modifications due to exhalation delay^39,43^. The restrictive pattern of Parkinson’s disease is characterized by rigidity, kyphosis, decreased thoracic compliance, intercostal muscle tremor, weakness, and fatigue from the continuous movement of the respiratory act, as well as abnormal respiratory muscle activity and unsynchronized contraction of these muscles, which affect breathing pattern. These impairments reduce values such as vital capacity and forced expiratory volume in one second (FEV1)^37,40–43^. On the other hand, a systematic review of McMahon, Blake and Lennon (2020) cited no correlation between restrictive dysfunction and rigidity, tremor or bradykinesia, although obstructive disorders commonly cite these characteristics as the main cause of it^44–46^.Because our sample consisted of PD patients with a modified Hoehn and Yahr score of 1.5 to 3.0 and mild to moderate symptoms, there was no prominent stooped posture that could effectively restrict air flow to the lungs, and normalized volumes were greater than 80%, which could explain the lack of a significant difference on dry needling intervention. Even with the TP in the trapezius muscle prior to the DNT session, some participants formally reported pain prior to the DNT session and improvement on this pain at the follow-up assessment; however, this was not indicative of volume improvement after the one DNT session.

Several limitations were found in this study. First, given the number of DNT sessions offered to each participant, which is only one, more sessions could be proposed for a more longitudinal study. However, we took into account the travel logistics of the people with PD, who were usually accompanied by a family member may alter the family’s routine activity. Second, given the severity of PD in the mild to moderate stages, proposing the DNT for PD patients in more advanced stages, where lung impairment may be more prominent, may yield positive results for the patient. Further studies using the dry needling technique in the population with PD are needed regarding trigger points, muscle spasms or stiffness, and the repercussions on thoracic and pulmonary volumes, as well as the possible biomechanical alterations to be considered. Finally, in addition to vital capacity and lung volume, additional clinical assessments such as the pain scale, chest expansion measurement, endurance test, and muscle flexibility test should be investigated.

## CONCLUSIONS

No statistically significant improvements in vital capacity measured by lung volumes were observed in people with PD after a session of the DNT to release trigger points in the trapezius. These findings can provide evidence that a single session of dry needling does not help to improve respiratory function in people with PD. However, our results showed a slight increase in the mean of maximum and of mean volumes of the patients who received the technique, which may be clinically relevant when referring to a progressive neurodegenerative disease. The more sessions of dry needling and complementary treatment that may need to be explored and observed to get the best treatment for respiratory function in people with PD.

## Data Availability

All data produced in the present study are available upon reasonable request to the authors.

## ACKNOWLEDGMENTS

The authors would like to thank Dr. Fuengfa Khobkhun (Mahidol University - Thailand), Prof. Jim Richards (University of Central Lancashire - United Kingdom) and patients of Hospital das Clínicas HC-FMRP/USP.

## AUTHOR CONTRIBUTIONS

Conceptualization, A.K.T. and P.R.P.S.; methodology, A.K.T., A.C.G. and P.R.P.S.; formal analysis, A.K.T. and P.R.P.S.; investigation, A.K.T.; A.G.C. and R.L.M.M. writing original draft preparation, A.K.T.; writingreview and editing, A.K.T.; A.C.G.; A.G.C.; R.L.M.M.; V.T. and P.R.P.S. visualization, A.K.T.; A.C.G.; A.G.C.; R.L.M.M.; V.T. and P.R.P.S.; supervision, A.C.G.; and P.R.P.S.; project administration, P.R.P.S.; All authors have read and agreed to the published version of the manuscript.

## FUNDING

This study was financed in part by the Coordenação de Aperfeiçoamento de Pessoal de Nível Superior - Brasil (CAPES) - Finance Code 001 and grant #2019/17729-0, #2010/20538-7, São Paulo Research Foundation (FAPESP).

## INSTITUTIONAL REVIEW

The study was conducted in accordance with the Declaration of Helsinki, and approved by the Institutional Review Board of the School of Physical Education and Sport of Ribeirão Preto at the University of São Paulo (process nž 45669720.2.0000.5659. April 5th, 2022) and Hospital of USP’s Medical School of Ribeirão Preto as as a co-participating institution (process nž 45669720.2.3001.5440. July 22, 2022)

## INFORMED CONSENT

Informed consent was obtained from all subjects involved in the study.

## DATA AVAILABILITY

Data can be made available upon request.

## CONFLICTS OF INTEREST

The authors declare no conflict of interest.

## ABBREVIATIONS

BMI: Body mass index
CI: Confidence interval
COPD: Cronic Obstrutive Pulmonary Disease
DN: Dry Needling group
DNT: Dry Needling Technique
BL: Baseline assessment
IA: Immediately after intervention assessemnt
FU: One-week follow-up
FEV1: Forced expiratory volume in one second
HR: Heart rate
HY: scale Modified Hoehn & Yahr scale
max_vol: Maximum volume
mean_vol: Mean volume
MM: Anova Mix model analysis of variance
PD: Parkinson’s Disease
ROM: Range of Motion
Sham: Sham needling group
SpO2: Partial Oxygen Saturation
TP: Trigger Point
UPDRS: Unified Parkinson’s Disease Rating Scale

## Notes

### Competing Interest Statement

The authors have declared no competing interest.

### Clinical Trial

RBR-4mg56yt

### Funding Statement

This study was financed in part by the Coordena(&ccedil)ao de Aperfei(&ccedil)oamento de Pessoal de Nivel Superior - Brasil (CAPES) - Finance225
Code 001 and grant #2019/17729-0, #2010/20538-7, Sao Paulo Research Foundation (FAPESP)

### Author Declarations

Institutional Review Board of the School of Physical Education and Sport of Ribeirao Preto at the University of Sao Paulo gave ethical approval for this work (45669720.2.0000.5659. April 5th, 2022) Institutional Review Board of Hospital of USP Medical School of Ribeirao Preto as co-participating institution gave ethical approval for this work(45669720.2.3001.5440.July 22, 2022)

